# Cognitive impairment is differentially associated with depression and anxiety at six-months post-stroke

**DOI:** 10.1101/2020.09.24.20200972

**Authors:** Owen A Williams, Nele Demeyere

## Abstract

**Objective:** This study investigated the associations between general cognitive impairment and domain specific cognitive impairment with post-stroke depression and anxiety at six-months post-stroke.

**Methods:** Participants were a subset of 437 stroke patients from the OCS-CARE study who were followed up at a 6 months post-stroke assessment. Depression and anxiety symptoms were assessed by the Hospital Anxiety and Depression Scale sub-scales. General cognitive impairment was assessed using the Montreal Cognitive Assessment and stroke-specific cognitive domain impairments was assessed using the Oxford Cognitive Screen. Multivariable linear regression was used to examine the associations between cognition and depression/anxiety symptoms, controlling for acute stroke severity and ADL impairment, age, sex, education, and co-occurring post-stroke depression/anxiety.

**Results:** Six-month post-stroke depression was associated with six-month impairment on the MoCA (beta [*b*] =0.96, standard error [*SE*] =0.31, *p*=0.006), and all individual domains assessed by the OCS including spatial attention (*b*=0.67, *SE*=0.33, *p* =0.041), executive function (*b*=1.37, *SE*=0.47, *p*=0.004), language processing (*b*=0.87, *SE*=0.38, *p*=0.028), memory (*b*=0.76, *SE*=0.37, *p*=0.040), number processing (*b*=1.13, *SE*=0.40, *p*=0.005), and praxis (*b*=1.16, *SE* =0.49, *p*=0.028). Post-stroke anxiety was associated with impairment on the MoCA (*b*=1.47, *SE*=0.42, *p*=0.001), and spatial attention (*b*=1.25, *SE*=0.45, *p*=0.006), but these associations did not remain significant after controlling for co-occurring post-stroke depression.

**Conclusion:** The different profiles of associations between cognitive impairment and post-stroke depression and anxiety suggest that cognitive impairment is highly related to depressive symptomatology, but associations observed between cognitive impairment and anxiety may actually be the result of co-morbid post stroke depression.

## Introduction

Mood disorders are common after stroke, occurring at higher rates than in the general population.^1, 2^ Post-stroke depression and anxiety can have a negative impact on long-term recovery.^3^ Cognitive status is an important predictor of post-stroke depression.^4^ However, research has provided inconsistent evidence of domain specific associations between cognition and post-stroke mood.

One of the most consistently reported risk factors for post-stroke depression is cognitive impairment.^4^ However, research into the impact of domain-specific cognitive impairment has yielded inconsistent findings. Acute impairments in language, visual memory, visuospatial neglect, executive functioning and working memory have been associated with depressive symptoms at three-months^5^ and six-months post-stroke.^6^ In comparison to post-stroke depression, the association of post-stroke anxiety with cognition has received very little attention. One study reported associations between post-stroke anxiety and impaired processing speed but not executive function or verbal memory.^7^ Anxiety at one-year post-stroke has been associated with overall cognition.^8^

The majority of studies have only measured acute/sub-acute cognitive impairment^5-7^ and it remains unclear how persistent cognitive impairment (> six-months post-stroke) is associated with mood disorders after six months post-stroke. Furthermore, despite their high comorbidity,^7^ depression and anxiety are rarely studied together^7, 8^ and models examining cognition-mood associations seldom control for co-occurrence of depression and anxiety. The aim of this study was to examine domain specific relationships between lasting cognitive impairment and risk of post-stroke depression and anxiety at six-months post-stroke while controlling for counterpart mood symptoms i.e. is cognitive impairment associated with depression independent of anxiety and vice-versa?

## Methods

### Participants

Participants were a subsample of the OCS-Care Study, a multi-centre sample of stroke patients recruited consecutively between July 2014 and July 2016 from 37 sites in England, UK.^9^ Originally, participants were included if they met the following criteria: (i) acute stroke patient (within 10 weeks of confirmed stroke); (ii) adult (≤90 years of age); (iii) able to concentrate sufficiently for one hour as judged by the multidisciplinary care team in the hospital; (iv) have sufficient language comprehension to pass the first orienting tests (the OCS Picture Naming and Semantics tasks); and (v) willing and able to give informed consent themselves. Further to this, participants were only included in the present analyses if they had complete data including stroke severity and activities of daily living during the acute stage and had completed cognition and mood assessments at six months follow-up. This was necessary to allow for comparison between statistical models for different cognitive domains as case-wise exclusion based on missing data may lead to slightly different samples being included in different models. All participants provided written informed consent. All procedures were in accordance with the Declaration of Helsinki and approved by the West Midlands – Coventry and Warwickshire National Research Ethical Committee (REC reference 12/WM/00335).

### Mood assessment

Depression and anxiety symptomatology were assessed using the Hospital Anxiety and Depression Scale (HADS).^10^ The HADS consists of seven items each for depression and anxiety with answers rated 0-3 for each question providing two subscales with a range of 0-21 for depression (HADS-D) and anxiety (HADS-A). Scores above seven indicate possible cases while scores above 11 indicate probable cases of depression/anxiety. Since its introduction, the HADS has been used extensively in clinical practice and research and has been found to have high sensitivity and reliability.^11^ The HADS was administered to participants during a 6 month face-to-face follow-up assessment. While we used the cut-off scores to describe the prevalence of depressive and anxious symptoms, the HADS subscales were entered into analyses as continuous variables.

### Cognitive assessment

General cognitive status and domain specific cognitive impairments were assessed in a face-to-face research visit either at home or in the clinic at 6-month post-stroke.

Cognitive status was assessed using the Montreal Cognitive Assessment (MoCA).^12^ The MoCA consists of a single A4 page and can be administered in approximately 15 minutes. While it contains assessments of several cognitive functions, there are no separable norm data to determine cut-offs in subsections. Instead, a total score out of 30 is calculated, with scores < 26 considered indicative of cognitive impairment.^12^ Although originally developed for mild cognitive impairment and dementia, the MoCA is commonly used to screen for cognitive impairment in stroke patients and has been shown to have good sensitivity to cognitive impairment after stroke.^13^

Domain specific impairments were assessed using the Oxford Cognitive Screen (OCS).^14^ The OCS is a cognitive screening tool designed specifically to assess common cognitive impairments in stroke patients. The test consists of 12 subtest scores that can be categorised into six cognitive domains: spatial attention (object and spatial neglect), executive function (trails switching accuracy), language (picture naming, semantic understanding, sentence reading), episodic memory (orientation, verbal recall, verbal recognition), number processing (number writing, calculation), and praxis (hand gesture imitation). Subtests are binarized into impaired/unimpaired based on normative scores for each subtest. Each cognitive domain is treated as impaired if at least one subtest in the domain is classed as impaired. A detailed description of the tests alongside normative data, and validation have been published previously.^14, 15^

### Acute stroke severity assessment

Baseline assessment of stroke severity was assessed vie the National Institute of Health Stroke Scale (NIHSS).^16^ The NIHSS is a brief 11-item observation scale that addresses cognitive and motor problems after stroke. Baseline stroke impact on daily life functioning was assessed via the Barthel Index of Activities of Daily Living (ADL). The Barthel Index consists of 10 items that measure a person’s daily functioning, specifically the activities of daily living and mobility.

### Statistical analysis

All analyses were carried out in R (version 3.6.3). Preliminary analyses to examine group differences in demographics, clinical stroke features, and cognitive impairment between participants included in the analysis and those excluded for missing data were conducted using univariate analyses (t-tests for continuous variables, χ2-test for categorical data). For the core analyses, pirate plots and univariate linear regression models were used to assess associations between cognitive metrics and HADS subscale scores.

We present two sets of analyses where the effects of cognition on depression or anxiety were examined separately. Multivariable linear regression was applied using the lm function in R to assess the association between cognition and HADS-D/HADS-A scores while controlling for confounding variables. Cognitive predictors included MoCA cognitive impairment (score < 26), OCS total number of impaired domains (categorised as 0, 1, or 2 or more domains impaired, due to a skewed distribution), and impairment on each individual cognitive domain separately. Two sequential models were applied for each cognitive measure as predictor variables with HADS-D/HADS-A score as the dependent variable. Model 1 controlled for age at stroke, sex, education (years), NIHSS, and Barthel score. To assess whether cognition is associated with depression independently of anxiety and vice versa, Model 2 additionally controlled for HADS-A in the models predicting HADS-D and HADS-D in the models predicting HADS-A. Several covariates were binarized due to skewed or limited range distributions and were classed as impaired vs spared based on clinical cut-offs. Stroke severity was classified as minor stroke vs major stroke with NIHSS scores > 3 indicating major stroke.^17^ The Barthel score was classified as impaired vs unimpaired daily functioning, with a score < 95 indicating impairment.^18^ Age at stroke and years of education were mean centred. The increased risk of Type I error due to multiple comparisons was corrected for by applying the false discovery rate (FDR) procedure as implemented using the p.adjust function in R.

In addition, logistic regression was applied as secondary analysis. Models with symptom group membership as the dependent variable based on HADS subscale cut-offs of eight of more to be considered depressive or anxious cases. Only cognitive variables that were found to be associated with HADS subscales in Model 2 were analysed. This was because there was a large proportion of participants who were both depressive and anxious. Thus, linear models were necessary first to interrogate the associations of interest while controlling for co-occurring mood symptoms. Covariates again included age at stroke, sex, education, NIHSS score, and Barthel score. Years of education was binarized into high (those with more than 11 years of education) and low (those who left school before age 16) due to the violation of the assumption of linearity between the logit of the outcome and each predictor variable.

## Results

In the original OCS-CARE study, 467 participants completed assessments and questionnaires during the acute stage post stroke and at a 6-month follow-up.^9^ Thirty of these participants had missing data for at least one variable of interest and were therefore excluded from all subsequent analysis in order for statistical models to be compared across the exact same sample. Table S1 shows that there were no significant differences in demographics or clinical stroke features between the subsample and the 30 excluded individuals. However, a greater proportion of the excluded participants did have an impairment in the language domain (33.3%) compared to the subsample with complete data (17.9%), χ2 = 3.886, p = 0.0487, suggesting aphasia symptoms may have contributed to increased missingness of questionnaire data in particular. In total, 437 participants were included in all analyses.

Table 1 shows the number of possible and probable cases for depression and anxiety in the sample at the six-month post stroke assessment (based on HADS cut offs). Of the participants who were non-cases, 266 (60.8% of whole group) participants did not have depression or anxiety symptoms. Of the possible and probable cases, 77 (17.6% of whole group) participants had both depression and anxiety symptoms.

**Table 1.**
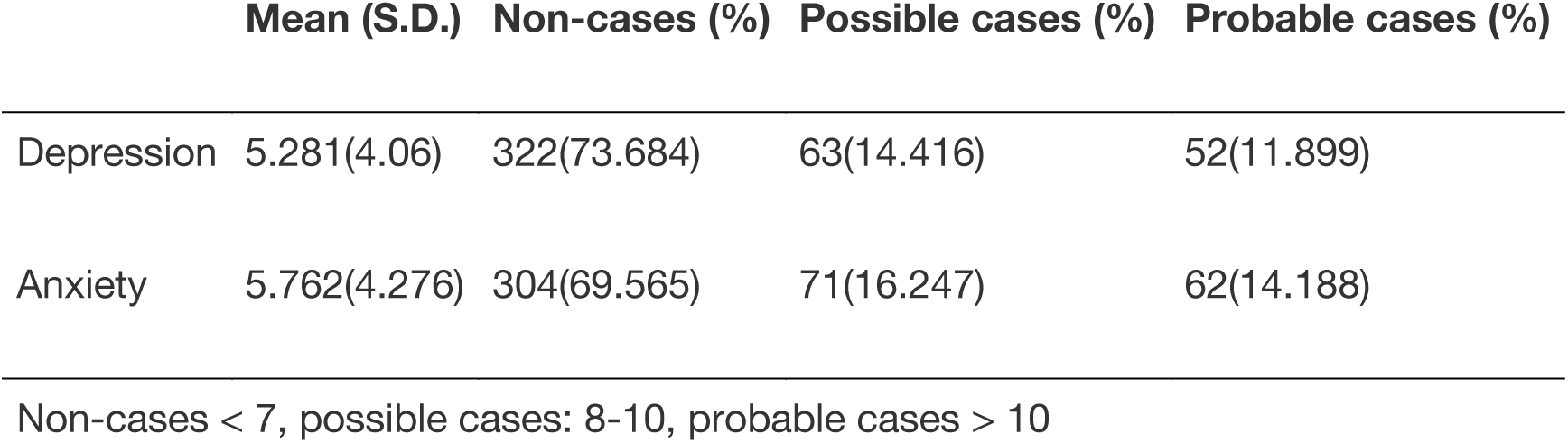
Rates of depression and anxiety at six-months post-stroke

|  | Mean (S.D.) | Non-cases (%) | Possible cases (%) | Probable cases (%) |
| --- | --- | --- | --- | --- |
| Depression | 5.281(4.06) | 322(73.684) | 63(14.416) | 52(11.899) |
| Anxiety | 5.762(4.276) | 304(69.565) | 71(16.247) | 62(14.188) |

Non-cases < 7, possible cases: 8-10, probable cases > 10

## Depression

Being impaired in the MoCA or having one, or two or more impairments in the OCS was associated with higher HADS-D scores compared to no impairments (see Figure 1). Supplementary Figure 1 shows pirate plots comparing HADS-D for individual cognitive domains on the OCS, all 6 domains showed significantly higher HADS-D scores in the impaired groups compared to not impaired groups (p <0.01).

**Figure 1:**
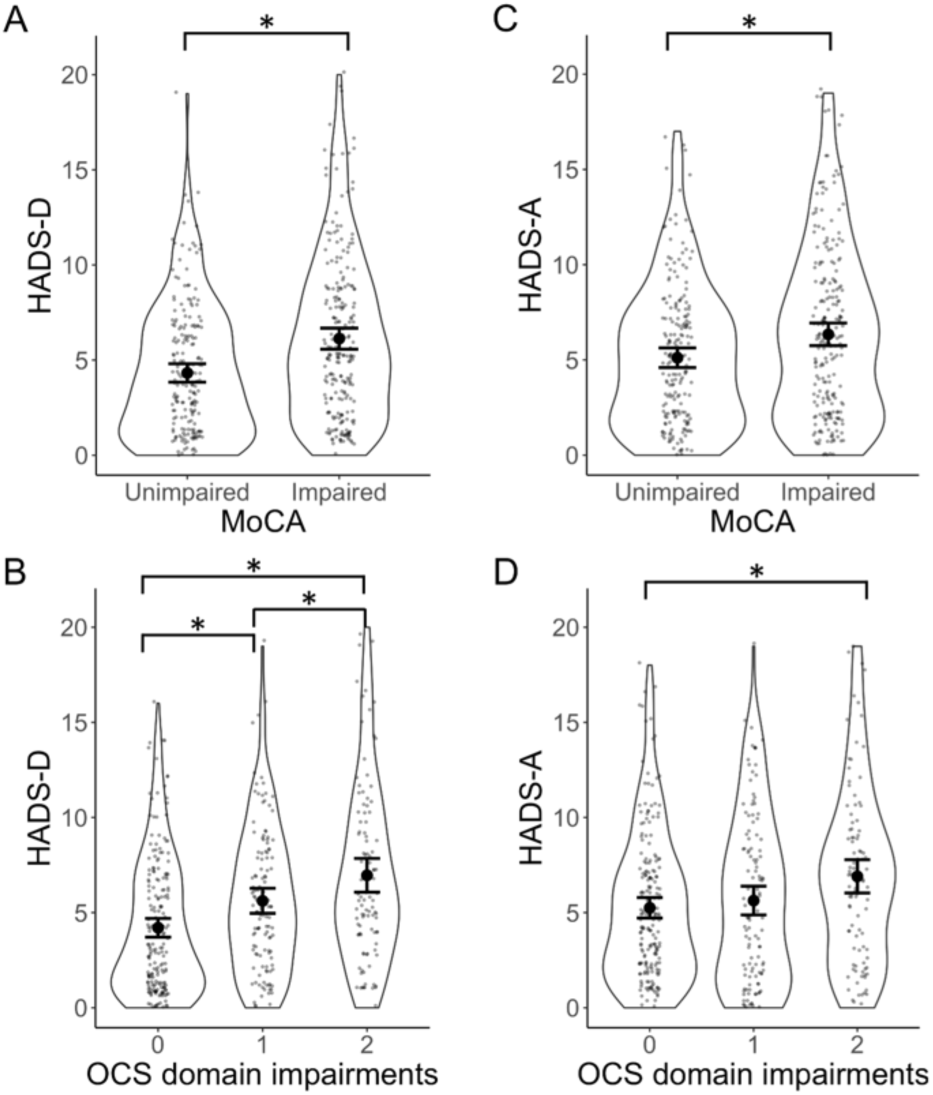
Pirate plots describing the distributions of depression and anxiety scores. a: HADS-D scores compared between unimpaired and impaired participants on the MoCA, b; HADS-D scores compared between participants with no cognitive domains impaired (0), one domain impairment (1), and two or more cognitive domain impairments (≥2). c: HADS-A scores compared between unimpaired and impaired participants on the MoCA, d; HADS-A scores compared between participants with no impairment, one impairment, and two or more impairments. Small dots indicate raw data, large dots indicate means with 95% confidence intervals, full data distributions are shown with the smoothed density curves. * p < 0.05

All cognitive metrics were positively associated with HADS-D after FDR correction for multiple comparisons. Having two or more domain impairments on the OCS was the predictor variable with the largest Beta value, suggesting that the accumulative effects of impairment on multiple domains is an important risk factor of depressive symptoms. Overall, the beta values were slightly attenuated in Model 2 which additionally controlled for anxiety symptoms using the HADS-A subscale (see Table 2).

**Table 2.**
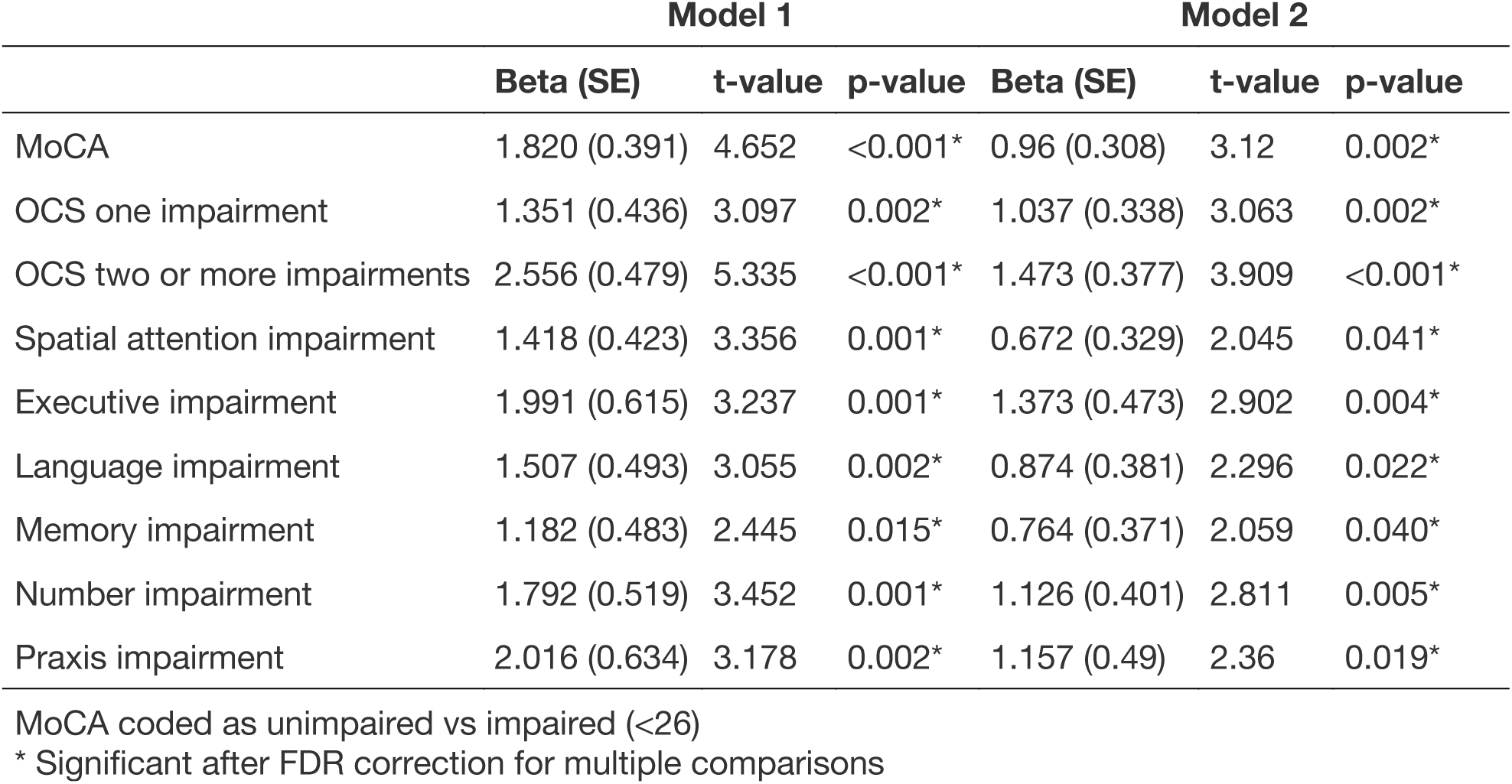
Effects of cognitive impairment on risk of depression at six months post stroke. Results from models 1 and 2

Results from the logistic regression carried out in secondary analyses, are presented in Figure 2. There was an increased risk of possible depression in individuals with an impairment on the MoCA (odds ratio [OR] = 3.04, 95% confidence interval [CI] = 1.84-5.14), one cognitive domain impairment on the OCS (OR= 2.19, CI = 1.25-3.87), two or more cognitive domain impairments on the OCS (OR = 3.37, CI= 1.85-6.23), impaired spatial attention (OR = 1.83, CI = 1.10-3.03), impaired language (OR = 1.80, CI = 1.02-3.16), impaired number processing)OR = 2.46, CI = 1.37-4.43) and impaired praxis (OR = 2.40, CI = 1.16-4.95). Only impairment on executive function (OR = 1.82, CI = 0.88-3.7) and memory (OR = 1.75, CI = 0.99-3.05) were not associated with increased risk of depression. Groupwise comparisons of the depressed vs. not depressed groups for logistic regression are shown in Supplementary Table S2.

**Figure 2:**
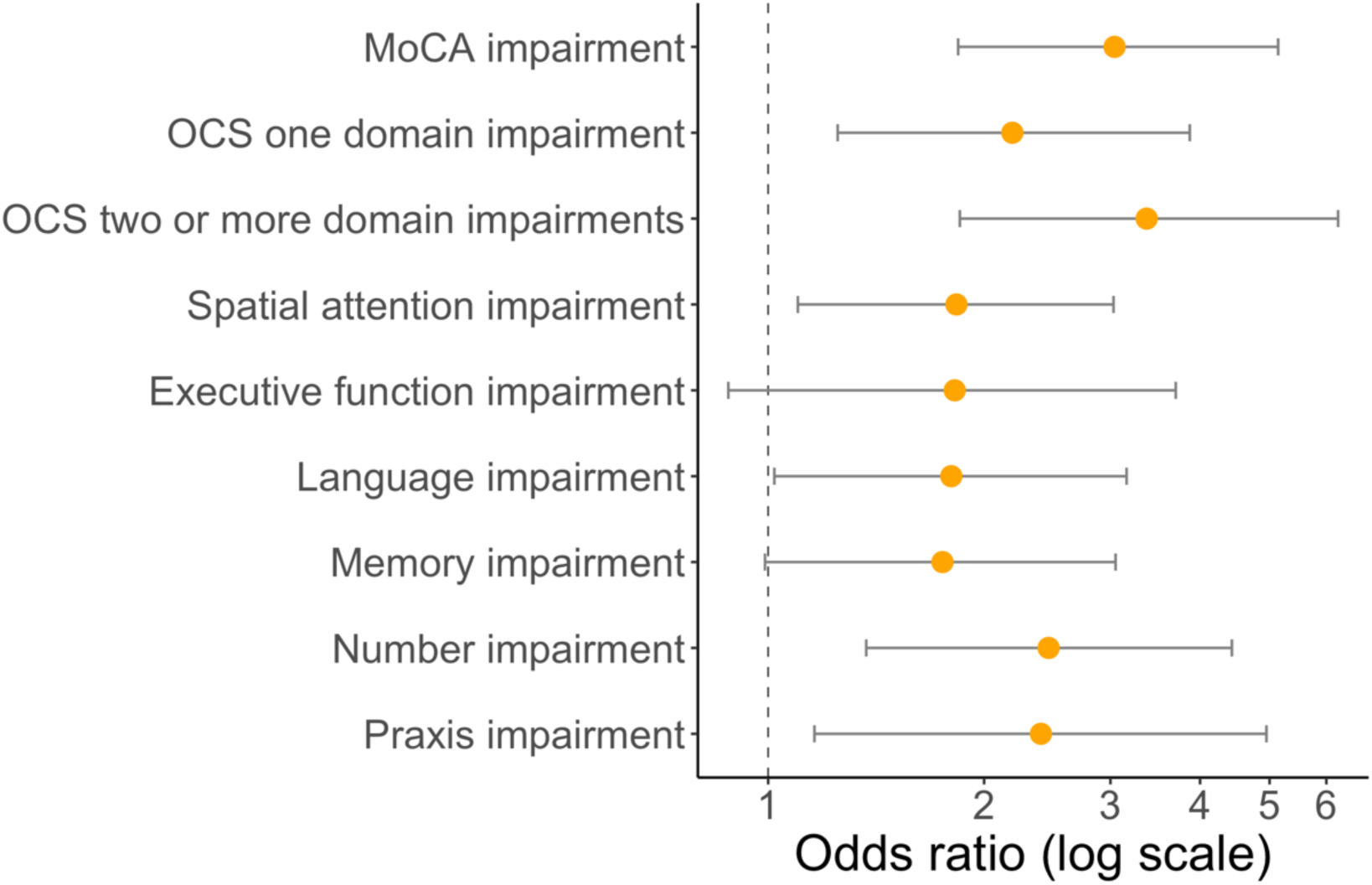
Odds ratios for risk of possible depression for each cognitive metric assessed. Bars indicate confidence intervals. Model is corrected for age, sex, education, NIHSS score, and Barthel score

### Anxiety

Being impaired in the MoCA or having two or more impairments in the OCS was associated with higher HADS-A scores compared to no impairments (see Figure 1). When comparing HADS-A for individual cognitive domains on the OCS, only impairments in spatial attention (p = 0.009), language (p = 0.042), and number processing (p = 0.026) showed higher HADS-A scores compared to individuals not impaired in those domains (Additional pirate plots for separate cognitive domains are provided in Supplementary Figure S1).

In Model 1 (controlling for age at stroke, sex, education, NIHSS score, and Barthel score) general cognitive impairment on the MoCA, having two or more cognitive domain impairments on the OCS, and impaired spatial attention were positively associated with HADS-A scores after FDR correction for multiple comparisons. Language impairment, number impairment, and praxis impairment were positively associated but did not survive FDR multiple comparison correction. See Table 3 for multivariable regression values with HADS-A scores as the dependent variable.

**Table 3.** Effects of cognitive impairment on risk of anxiety at six months post stroke. Results from models 1 and 2

|  | Model 1 |  |  | Model 2 |  |  |
| --- | --- | --- | --- | --- | --- | --- |
|  | Beta (SE) | t-value | p-value | Beta (SE) | t-value | p-value |
| MoCA | 1.465 (0.421) | 3.477 | 0.001* | 0.225 (0.335) | 0.671 | 0.502 |
| OCS one impairment | 0.539 (0.473) | 1.138 | 0.256 | -0.39 (0.371) | -1.051 | 0.294 |
| OCS two or more impairments | 1.858 (0.520) | 3.571 | <0.000* | 0.101 (0.416) | 0.242 | 0.809 |
| Spatial attention impairment | 1.251 (0.452) | 2.767 | 0.006* | 0.284 (0.353) | 0.803 | 0.422 |
| Executive impairment | 1.034 (0.662) | 1.563 | 0.119 | -0.342 (0.513) | -0.667 | 0.505 |
| Language impairment | 1.058 (0.529) | 2.001 | 0.046 | 0.023 (0.411) | 0.055 | 0.956 |
| Memory impairment | 0.695 (0.517) | 1.343 | 0.180 | -0.119 (0.399) | -0.298 | 0.766 |
| Number impairment | 1.116 (0.558) | 2.000 | 0.046 | -0.119 (0.435) | -0.273 | 0.785 |
| Praxis impairment | 1.438 (0.68) | 2.114 | 0.035 | 0.054 (0.529) | 0.102 | 0.919 |

MoCA coded as unimpaired vs impaired (<26)
\* Significant after FDR correction for multiple comparisons

As no cognitive metrics were associated with HADS-A independent of depression, no logistic regression models were carried out for anxiety.

## Discussion

In a large sample of stroke patients who were followed up at six-months post-stroke, we found that a general measure of cognitive impairment (MoCA score standard cut off) as well as impairment in multiple domains, as measured by the OCS, was associated with elevated levels of post-stroke depressive symptomatology independent of stroke severity, ADL impairment, age at stroke, sex, education and co-occurring anxiety symptomatology. Post-stroke anxiety showed limited associations with cognition and these were lost when controlling for co-occurring post-stroke depressive symptomatology. These results suggest that while overall severity of cognitive impairment shows similar associated risk for post-stroke depression or anxiety, there are different patterns of associations between specific cognitive impairments and risk of post-stroke depression compared to post-stroke anxiety and that observed associations between post-stroke cognitive impairment and anxiety may in fact be due to co-occurring depression. The finding that these associations were found to be independent of stroke severity and initial disability has important implications for targeted therapy of post-stroke cognitive impairment and mood disorders.

The results from the fully adjusted Model 2 for depression confirm previous reports that general cognitive impairment after stroke is associated with depression at six-months post-stroke.^6, 19^ By using the stroke specific OCS assessment, this study extended these findings to show that having an impairment in one or more cognitive domains increases depressive symptomatology, suggesting that there is an additive effect of domain specific cognitive impairments on the risk of depressive symptomatology, with those having more than two cognitive domain impairments at greater risk of post-stroke depression. Furthermore, we were able to assess associations between impairment in specific domains and depressive symptomatology. We found positive associations between depressive symptomatology and impairments in all domains assessed, including spatial attention, executive function, language processing, memory, number processing and praxis. While spatial attention (including neglect), executive functioning, language processing and memory have been more commonly studied and found to be related to post-stroke depression,^5, 6, 20^ number processing and praxis have not received the same level of investigation. Here we show that these domains are important contributors in the profile of cognitive impairment related to post-stroke depression as they each have the highest odds ratios of the six individual domains assessed by binary logistic regression in the secondary analyses. Furthermore, impairments in these two domains are common among stroke survivors.^21^

Episodic memory as assessed by the OCS was significantly associated with post-stroke depressive symptomatology. This is in contrast with an earlier report from Hosking et al.^22^ that episodic verbal memory as assessed by the Verbal Paired Associates subtest of the Wechsler Memory Scale-Revised^23^ was not associated with post-stroke depression. As the OCS was designed specifically for stroke patients and allows for non-verbal responses when oral communication is impaired, it is possible that it provides a more accurate description of memory impairment compared to the WMS-R subtests in stroke patients and that may explain the discrepancy in our current findings. While Nys et al.^6^ reported significant associations between visual memory and 3-month PSD symptomatology, they did not find any association with verbal memory impairment, again suggesting that relying on verbal abilities in acute stroke patients may cloud associations between episodic memory and PSD. Hommel et al.^5^ did find significant associations between acute impairments in verbal episodic memory and three-month post-stroke depression, however the association was lost when they controlled for demographics and clinical stroke features.

Our finding that executive function was associated with post-stroke depressive symptomatology in linear regression models but was not associated with post-stroke depression in the logistic regression was surprising. In similar logistic regression analyses, positive associations between post-stroke depression symptoms and impairments in various aspects of executive function have been consistently reported.^5, 6, 22^ Executive function in the OCS is assessed using a trails task which involves set shifting and inhibiting previously learned responses. In contrast to many executive function tasks, impairment is judged solely on accuracy, with no time component taken into account due to the high proportion of stroke patients with motor impairments. Although other executive function metrics are often corrected for speed (e.g. Trails-B – Trails-A^24^), it is still possible that these methodological differences may contribute to different profiles of executive function impairments. Trails-B has been associated with post-stroke depression but this association did not remain when demographics and clinical features were controlled for. However, in the same study, performance on the Hanoi Tower^25^ was associated with post-stroke depression independently of demographics and clinical stroke features. This is a complex task that measures multiple facets of executive function and may be more sensitive to deficits than the OCS trails task. Nys et al. administered a comprehensive battery of executive function tests that were shown to be associated with post-stroke depression. However, the models did not control for stoke severity or other clinical features.^6^

For post-stroke anxiety, we found that general cognitive impairment and severity as measured by number of domains impaired on the OCS were related to increased anxiety symptoms. This is supported by a previous finding of a cross-sectional relationship between post stroke anxiety and general cognition at one-year post-stroke.^8^ However, others have reported no associations between anxiety and cognition^26^ When individual components of cognition were analysed, only impairment on the praxis task was significantly associated with anxiety symptoms, independently of other important risk factors. To our knowledge this is the first time that apraxia has been found to be associated with post stroke anxiety. The fact that no other domain tested was associated with post-stroke anxiety is similar to previously reported findings that executive function and verbal memory were not related to anxiety at three-months post-stroke.^7^ However, when depressive symptoms were added in Model 2, there were no significant associations between anxiety symptoms and any of the cognitive measures used. Very few studies have investigated post-stroke depression and anxiety together. Barker-Collo^26^ had previously reported that performance on cognitive tests explained more variance in post stroke depressive symptomatology that it did in anxiety symptomatology but the study did not control for co-occurrence. As post-stroke depression and anxiety are highly correlated,^8^ it is important to control for depressive symptoms when investigating independent associations between cognitive impairment and post stroke anxiety. Lee et al.^8^ found that while acute impairment on the mini-mental state exam was associated with anxiety in the first two-weeks after stroke, this effect was lost once controlling for depressive symptoms. At one-year post stroke follow-up they found that MMSE scores were associated with anxiety independently of depressive symptoms present during the acute stage but did not control for co-occurring post-stroke depression at the one-year assessment. With 58% of participants who tested positive for possible/probable anxiety also testing positive for depression in this sample at six-months, we saw that controlling for depressive symptoms completely removed significant associations between cognitive impairment and anxiety. Therefore, it is possible that observed associations between post-stroke cognitive impairment and anxiety were actually being driven by associations with depression.

The results of this study should be considered in light of several limitations. While the results suggest that persistent cognitive impairment is associated with greater chance of stroke patients having symptoms of post stroke depression and to some extent also anxiety, the cross-sectional design does not allow for inference of causality. It may be the case that altered mood hinders the recovery of cognitive functioning or results in poorer performance on cognitive tasks. Support for the first idea comes from evidence that antidepressants have improved cognition after stroke.^27^ However, Nys et al.^6^ found that cognitive impairment during the acute stroke stage was associated with depressive symptomatology after six months post-stroke and more recent evidence has suggested that general cognitive impairment at six months post-stroke is associated with increased risk of depression five years post-stroke.^28^ It is possible that there is a bidirectional relationship between depression/anxiety and cognitive impairment in post-stroke individuals. Future longitudinal studies with multiple assessments of both cognition and mood would allow for a more advanced understanding of these relationships through, for example, random-intercept cross-lagged panel modelling.^29, 30^ It should be noted that there is some evidence that lesion location may be related to depression, with left hemisphere lesions conferring higher risk of post-stroke depression, which fits with reports of language impairments relating to higher rates of depression post stroke.^20^ Unfortunately, there was a large amount of missing data regarding the location of the stroke lesion in this sample, though the findings of in particular apraxia, number and language impairments associations with post-stroke depression fit with this. We suggest that the relationship with lesion location is likely being driven by the cognitive impairments, rather than any particular neuroanatomical reason for depressive symptomatology being related to the left hemisphere. As a result of the large amount of missing data, lesion location could not be included in our multivariable analyses. Pre-stroke depression has also been shown to be an important predictor of post-stroke depression.^28^ Unfortunately, such data was not available for the current dataset. Future work examining cognition-mood associations should examine interactions with pre-stroke depression and anxiety.

A key strength of this study is the use of the OCS to assess impairment in individual cognitive domains that are commonly affected in stroke and how they are associated with post-stroke depression and anxiety. The advantages of the OCS are that it was designed specifically to assess for common domain-specific cognitive impairments post-stroke, such as aphasia, apraxia, neglect etc whilst allowing for participants to complete assessments even when motor or language impairments are present.^15^ This means that some of the more severely impaired participants could continue to participate. However, it is clear from comparisons with the 30 participants that were excluded, that some with severe language impairments were still not able to complete all the necessary assessments to be included in the present analyses. Therefore, it is possible that by excluding some of the most severely aphasic patients, we are still underestimating the impact of post-stroke language impairment on depression/anxiety, despite this, a large sample of stroke patients contributed to this study providing substantial statistical power for analyses.

This is one of few studies that have examined post stroke depression and anxiety alongside wide ranging cognitive screening in the same cohort. We found different profiles of associations with domain specific cognitive impairment and depressive vs anxiety symptomatology at six-months post stroke. While post-stroke depression was associated with impairments in all cognitive domains tested, anxiety was not found to be associated with cognitive impairment independently of depressive symptoms. Future work should focus on longitudinal experiments in order to elucidate the temporal relationships between domain specific cognitive impairment and mood disorders in stroke survivors as these associations may well have important implications for therapeutic applications.

## Data Availability

Data can be accuired via email request to the author.

## Acknowledgements

The authors would like the thank the participants who to part in the OCS-CARE study and all research staff who contributed towards data collection.

